# Clinical Utility of SARS-CoV-2 Whole Genome Sequencing in Deciphering Source of Infection

**DOI:** 10.1101/2020.05.21.20107599

**Authors:** Toshiki Takenouchi, Yuka W. Iwasaki, Sei Harada, Hirotsugu Ishizu, Yoshifumi Uwamino, Shunsuke Uno, Asami Osada, Naoki Hasegawa, Mitsuru Murata, Toru Takebayashi, Koichi Fukunaga, Hideyuki Saya, Yuko Kitagawa, Masayuki Amagai, Haruhiko Siomi, Kenjiro Kosaki, Keio Donner Project

**Author notes:** These authors contributed equally to this work (Toshiki Takenouchi, Yuka W. Iwasaki). Correspondence to Kenjiro Kosaki, M.D. and Haruhiko Siomi, Ph.D. Keio University School of Medicine, 35 Shinanomachi, Shinjuku-ku, Tokyo, 160-8582, Japan, and.

## Abstract

The novel coronavirus disease (COVID-19) pandemic caused by SARS-CoV-2 is a major threat to humans. Recently, we encountered two seemingly separate COVID-19 clusters in a tertiary care medical center. Whole viral genome sequencing detected the haplotype of the SARS-CoV-2 genome and the two clusters were successfully distinguished by the viral genome haplotype. Concurrently, there were nine COVID-19 patients clinically unlinked to clusters #1 or #2 that necessitated the determination of the source of infection. Such patients had similar haplotypes to those in cluster #2 but were devoid of two rare mutations characteristic to cluster #2. This suggested that these nine cases of “probable community infection” indeed had community infection and were not derived from cluster #2. Whole viral genome sequencing of SARS-CoV-2 is a powerful measure not only for monitoring the global trend of SARS-CoV-2 but also for identifying the source of infection of COVID-19 at a level of institution.

## Introduction

The current coronavirus disease 2019 (COVID-19) pandemic caused by human severe acute respiratory syndrome coronavirus 2 (SARS-CoV-2) is a major worldwide medical problem. Since the emergence of COVID-19 patients is about to collapse health care system in many countries, the development of effective diagnostic and preventive measures is urgently needed.

SARS-CoV-2 belongs to single-strand RNA virus and has a rapid pace of mutagenesis (~ two new mutations per month). Currently, global monitoring of SARS-CoV-2 mutation dynamics is publicly available by Global initiative on sharing all influenza data (GISAID: https://www.gisaid.org/) (l)and Nextstrain (https://nextstrain.org/X2). These data are useful in tracing longitudinal and global trend of viral genome changes, however, their roles on preventive measures at an institutional level remain to be explored.

## Results

**Cluster #1:** the index patient was transferred from a local hospital to Keio University Hospital for the purpose of surgery of on 3/19/2020. The patient did not have any respiratory symptoms. On 3/23/2020, it was disclosed that the local hospital had had nosocomial infection of COVID-19 before the patient transfer. On the following day, this patient was found to be positive on the clinical reverse-transcription polymerase chain reaction (RT-PCR) testing (3). Subsequently, three healthcare workers and four additional patients on the same floor became positive on the clinical RT-PCR testing.

**Cluster** #2-Another cluster occurred among interns and junior residents during the last week of March 2020. All 99 interns and junior residents underwent the clinical RT-PCR testing and 20 were tested positive. The source of infection remained unknown in this cluster.

**Other patients unlinked to clusters #1 or** *#*2-Concurrently with the clusters #1 and #2, there were ongoing cases of newly diagnosed COVID-19 patients that were unlinked to clusters #1 or #2. This group was considered having “probable community infection.”

The nasopharyngeal samples from four subjects in cluster #1, 12 subjects with positive RT-PCR results of cluster #2 and samples from nine subjects from “probable community infection” underwent whole viral genome sequencing (see methods).

### Phylogenetic tree analysis showed concordance between epidemiological contact tracing and viral genome haplotype

The phylogenetic tree analysis was performed locally using the Augur program available from Nextstrain (2). The dataset included sequencing data of all 25 subjects, global dataset of GISAID (1) submitted by 2/29/2020 and all available Japanese data excluding those obtained from the cruise ship, *the Diamond Princess* (4). The data points from cluster #1 were distinct from data points from cluster #2 (Figure 1). The cluster #1 appeared to be derived from the original SARS-CoV-2 descent in the Wuhan at a relatively early stage, whereas data points from other Japanese COVID-19 cases including those in cluster #2 and “probable community infection” cases were clustered rather closely.

**Figure 1.**
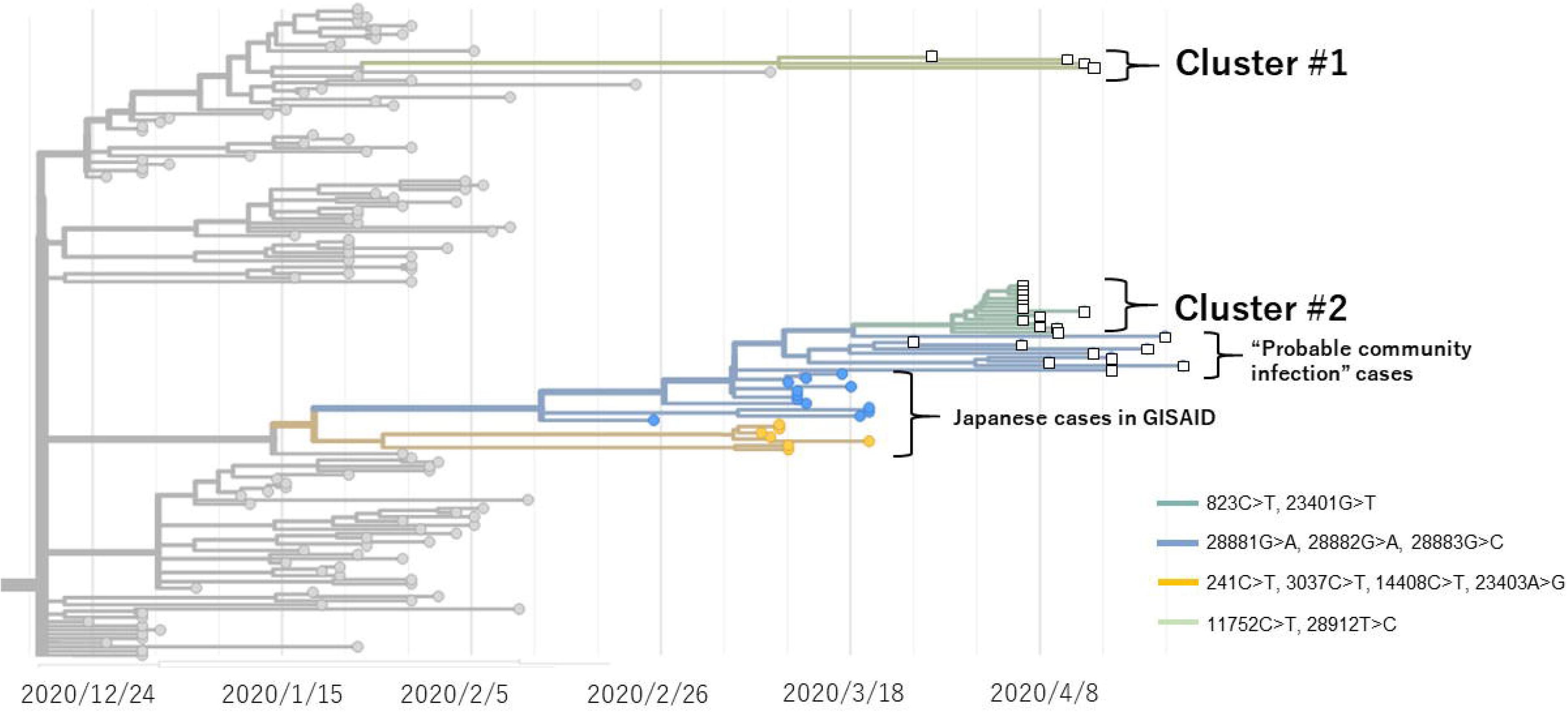
Result of phylogenetic tree analysis. Dots represent publicly available data points. Squares represents cases in the present study. Clades were defined following the color code at right bottom. Note that data points from cluster #1 and cluster #2 were distinct. Cluster #2, “probable community infection” cases and Japanese cases in GISAID formed clusters belong to a close branch (see Figure 2).

The viral genome haplotype analysis confirmed that cluster #1 and cluster #2 were distinguished by ten mutations (Figure 2). 11752C>T, 25665C>T, 26447C>T, 27700-27702delATT and 28912T>C were specific to cluster #1, whereas 241C>T, 313C>T, 3037C>T, 14408C>T, 23403A>G, 28881G>A, 28882G>A and 28883G>C were specific to cluster #2. The mutually exclusive haplotypes of clusters #1 and #2 provided molecular evidence that clusters #1 and #2 were caused by two different SARS-CoV-2 strains, thus were independent of each other. This was compatible with the results of in-hospital surveillance using the contact tracing.

**Figure 2.**
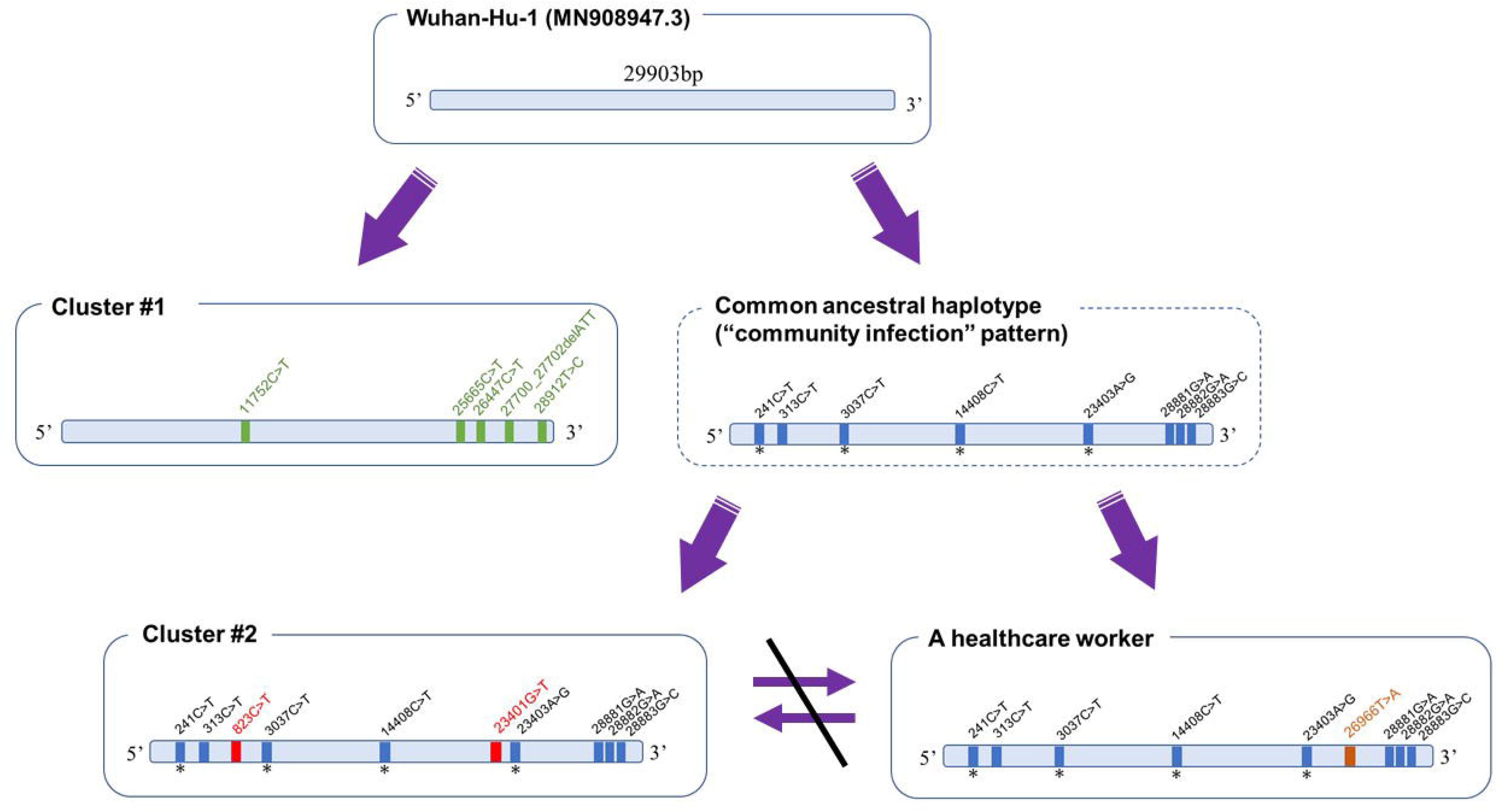
Postulated local evolutional history based on viral genome haplotype. Asterisks indicate common mutations that were present in approximately 56% of publicly available 8,604 SARS-CoV-2 cases downloaded from GISAID database as of April 16, 2020. Note that five mutations, i.e., 11752C>T, 25665C>T, 26447C>T, 27700_27702delATT and 28912T>C, were exclusively present in cluster #1, and was absent in cluster #2. Conversely, ten mutations, i.e., 241C>T, 313C>T, 823C>T, 3037C>T, 14408C>T, 23401G>T, 23403A>G, 28881G>A, 28882G>A and 28883G>C, were exclusively present in all 12 subjects from cluster #2, whereas none of such changes were present in those from cluster #1. Haplotypes of cluster #2 and all cases in “probable community infection” shared eight mutations, i.e., 241C>T, 313C>T, 3037C>T, 14408C>T, 23403A>G, 28881G>A, 28882G>A and 28883G>C, except for a pair of mutations of 823C>T and 23401G>T. A representative case of a healthcare worker at right bottom did not have either 823C>T or 23401G>T, but had 26966T>A, which was not present either in cluster #1 or cluster #2. This observation indicated that the strains carried by this healthcare worker and cluster #2 derived from the shared ancestor, i.e., “community infection pattern”, rather than direct cross-infection with each other.

### Viral genome haplotype of “probable community infection” cases

During the period of the two in-hospital clusters, there were nine cases of “probable community infection” that necessitated urgent determination of the source of infection. Although these nine subjects had a similar haplotype to that in cluster #2, there was a distinctive viral genomic signature: a pair of mutations, i.e., 823C>T and 23401G>T, was only present in those in cluster #2, but not in any of the “probable community infection” cases. This suggested that “probable community infection” cases indeed had community infection and was not derived from cluster #2, on the ground that it is unlikely for the mutations of the viral genome to revert spontaneously. It is most likely that cluster #2 and “probable community infection” cases derived from a common ancestral haplotype with eight mutations, i.e., 241C>T, 313C>T, 30370T, 144080T, 23403A>G, 28881G>A, 28882G>A and 28883G>C (Figure 2) that was presumably present in the neighborhood.

The utility of viral haplotype analysis was best exemplified by one healthcare worker in the “probable community infection” group. She worked as a full-time employee at Keio University Hospital. In addition, until 3/17/2020, she had an outpatient clinic once a week at the local hospital that was the origin of cluster #1. She had fever and became positive on the clinical RT-PCR on 4/13/2020. She denied any contacts with individuals from cluster #1 or #2. Her viral genome haplotype proved that she did not became infected with SARS-CoV-2 in the local hospital or Keio University Hospital.

## Discussion

In the present study, a rapid whole viral genome sequencing of SARS-CoV-2 on site successfully demonstrated distinctive viral genomic haplotype that were concordant with epidemiologic contact history in two intrahospital clusters. The viral genome haplotype was of use in deciphering the source of infection in COVID-19 cases that have no contact history to the existing COVID-19 clusters in a hospital.

From a standpoint of infection control in a hospital, whole genome viral sequencing helps determine whether newly diagnosed patients have nosocomial infection or community infection. In the situation of nosocomial infection, in addition to isolation of confirmed positive cases, a thorough contact tracing to search undiagnosed COVID-19 healthcare workers and inpatients is essential. In contrast, in the situation of community infection, such a thorough intrahospital surveillance is not necessary. Epidemiologic contact tracing combined with viral genomic data would be key for effective preventive measures against COVID-19.

The acquisition of whole viral genome sequence has implications in the future basic research perspective. In the present study, the whole viral genome sequencing not only provided nucleotide signatures of SARS-CoV-2 strains, but also identified 39 different mutations with 22 being amino acid substitutions in 25 samples. Research and development of vaccine and antibodies targeting SARS-CoV-2 should be pursued in view of this variability of viral protein sequences.

## Methods

PCR based amplification was performed using the Artie ncov-2019 primers, version 3 (https://github.com/artic-network/artic-ncov2019/blob/master/primer_schemes/nCoV-2019/V3/nCoV-2019.tsv) in two multiplex reactions according to the globally accepted “nCoV-2019 sequencing protocol” (https://www.protocols.io/view/ncov-2019-sequencing-protocol-bbmuik6w). Sequencing library for amplicon sequencing was prepared using the Next Ultra II DNA Library Prep Kit for Illumina (New England Biolabs). Paired-end sequencing was performed on the MiSeq platform (Illumina, CA). The bioinformatic pipeline used in this study, “Variant calling pipeline for amplicon-based sequencing of the SARS-CoV-2 viral genome”, is available at https://cmg.med.keio.ac.ip/sars-cov-2/.

## Data Availability

We uploaded the full nucleotide sequences of our cohort to the GISAID database (https://www.gisaid.org/).

## Acknowledgements and Data Sharing

We downloaded the full nucleotide sequences of the SARS-CoV-2 genomes from the GISAID database (https://www.gisaid.org/). A table of the contributors is available in Acknowledgement Table. We uploaded the full nucleotide sequences of our cohort to the GISAID database. We thank all patients and healthcare workers who fought against COVID-19. This work was supported by Keio Donner Project and is devoted to the late Professor Shibasaburo Kitasato, the founder of Keio University School of Medicine.

## References

1. S. Elbe, G. Buckland-Merrett, Data, disease and diplomacy: GISAID’s innovative contribution to global health. Glob Chall1, 33-46 (2017).

2. J. Hadfield et al., Nextstrain: real-time tracking of pathogen evolution. Bioinformatics 34, 4121-4123 (2018).

3. K. Shirato et al, Development of Genetic Diagnostic Methods for Novel Coronavirus 2019 (nCoV-2019) in Japan. Jpn J Infect Dis 10.7883/yoken.JJID.2020.061 (2020).

4. L. F. Moriarty et al., Public Health Responses to COVID-19 Outbreaks on Cruise Ships - Worldwide, February-March 2020. MMWR Morb Mortal Wkly Rep 69, 347–352 (2020).Cluster #2

